# Globally elevated excitation-inhibition ratio in children with autism spectrum disorder and below-average intelligence

**DOI:** 10.1101/2021.11.10.21266171

**Authors:** Viktoriya O. Manyukhina, Andrey O. Prokofyev, Ilia A. Galuta, Dzerassa E. Goiaeva, Tatiana S. Obukhova, Justin F. Schneiderman, Dmitry I. Altukhov, Tatiana A. Stroganova, Elena V. Orekhova

**Affiliations:** Center for Neurocognitive Research (MEG Center), Moscow State University of Psychology and Education, Moscow, Russian Federation; Department of Psychology, National Research University Higher School of Economics, Moscow, Russian Federation; MedTech West and the Institute of Neuroscience and Physiology, Sahlgrenska Academy, The University of Gothenburg, Gothenburg, Sweden

**Keywords:** Autism Spectrum Disorders (ASD), Intelligence, Magnetoencephalography, Power spectrum, 1/f Power law, Excitation-inhibition balance

## Abstract

**BACKGROUND:** An altered balance of neuronal excitation and inhibition (E-I balance) might be implicated in the co-occurrence of autism and intellectual disability, but this hypothesis has never been tested. E-I balance changes can be estimated from the spectral slope of the aperiodic 1/f neural activity. Herein, we used magnetoencephalography (MEG) to test whether the 1/f slope would differentiate ASD children with and without intellectual disability.

**METHODS:** MEG was recorded at rest with eyes open/closed in 49 boys with ASD aged 6-15 years with a broad range of IQs, and in 49 age-matched typically developing (TD) boys. The cortical source activity was estimated using the LCMV beamformer approach. We then extracted the 1/f slope by fitting a linear function in to the log-log-scale power spectra in the high-frequency range.

**RESULTS:** The grand averaged 1/f slope was steeper in the eyes closed than in the eyes open condition, but had high rank-order stability between them. In line with the previous research, the slope flattened with age. Children with ASD and below-average (<85) IQ had flatter slopes than either TD or ASD children with average IQ. These group differences could not be explained by differences in signal-to-noise ratio or periodic (alpha and beta) activity.

**CONCLUSIONS:** The atypically flattened spectral slope of aperiodic activity in children with ASD and below-average IQ suggests a shift of the global E-I balance toward hyper-excitation. The spectral slope can provide an accessible non-invasive biomarker of the E-I ratio for translational research and making objective judgments about treatment effectiveness.

## BACKGROUND

The hypothesis of an altered ratio between neural excitation and inhibition (E-I ratio) as a probable cause of autism was first introduced by Rubenstein and Merzenich in 2003 (1) and received numerous confirmations in genetic and animal research (2). Indeed, the majority of the animal models of ASD target functioning of the E or I neurons and alter the balance of their activity (3, 4), while pharmacological drugs that correct this balance can help to rescue the disease phenotype, at least in experimental animals (4). Although Rubenstein and Merzenich initially related ASD to an elevated E-I ratio, later it became clear that some forms of ASD can be characterized by decreased E-I ratio (5, 6) and that alternations in the E-I ratio can be region-specific, reflecting homeostatic regulation of local E-I imbalances (7). Still, there is evidence that, in many cases, the global deficit associated with ASD can be characterized as predominant neuronal hyper- (8-10) or hypo- (5, 6) excitability. Therefore, despite simplification, the concept of a global E-I balance is important for understanding the pathophysiological mechanisms associated with ASD and for identifying appropriate treatments (2).

The genes that lead to the E-I imbalance through their effect on neurotransmission are often implicated in both ASD and intellectual disability (ID) (11-13). People with ASD and comorbid intellectual disability are more likely to have epilepsy than autistic individuals without intellectual disability (14, 15), possibly because of more severe disturbances of the E-I balance. Although such patients may benefit most from targeted pharmacological treatment, neuroimaging studies looking for ‘autism biomarkers’ rarely include individuals from the lower end of the IQ range (16).

Physiological parameters that quantify the degree and direction of the E-I imbalance in individuals with ASD – particularly in those with below-average IQ – could provide valuable biomarkers for translational research, stratifying patients for clinical trials, and making objective judgments about treatment efficacy (2).

Magnetoencephalography (MEG) results directly reflect the activity of populations of cortical neurons and could potentially help to estimate the E-I balance globally as well as in distinct cortical regions. The power spectrum of electrophysiological signals measured with MEG or EEG comprises the ‘periodic’ part that reflects rhythmic brain activity and the ‘aperiodic’ part – the ‘neural noise’ (17, 18). The power of the aperiodic component decreases exponentially with increasing frequency, which is reflected in a linear relationship between them on a logarithmic scale. The slope of this linear function becomes flatter (less negative) during maturation (19-21), reflecting the increasing complexity of neural networks and/or changes in their structure and neurochemistry. At the same time, the spectral slope is sensitive to changes in excitatory and inhibitory neurotransmission during low arousal states, e.g. anesthesia and sleep (22-26) and may reflect E-I imbalance in neuropsychiatric disorders (27). It has been argued that the flattened spectral slope may reflect an elevated background rate of neurons’ firing, which is decoupled from an oscillatory carrier frequency and is driven by an increased E-I ratio (i.e., ‘noise’) (18, 23).

Although a spectrum of resting-state neural activity can be separated computationally into periodic and aperiodic components, the estimation of the aperiodic 1/f slope in the presence of oscillatory activity may vary depending on the method used, frequency bandwidth chosen, etc (25, 28, 29). To overcome these obstacles, the 1/f spectral slope can be estimated in ∼30-70 Hz range, where rhythmic activity is usually absent (23, 25).

Here, we used MEG and individual MRI-based brain models to capture the spectral slope from high-frequency brain activity measured ‘at rest’ in boys with ASD and below-average IQ, those with average IQs, and in age-matched TD boys. We predicted that children with ASD and low IQ would yield flatter slopes of the aperiodic component of the power spectra than the other two groups of children, indicative of their globally disturbed (predominantly elevated) E-I ratios.

## METHODS AND MATERIALS

In addition to this section, there is an extensive methodological Supplemental Materials, which describes each aspect of the analysis in more detail.

### Participants

The participants were 49 boys with ‘non-syndromic’ ASD and 49 TD boys aged 6-15 years. The study has been approved by the Ethical Committee of the Moscow State University of Psychology and Education. Verbal assent to participate in the experiment was obtained from all the subjects; parents/caregivers provided informed written consent.

For all these participants the MEG data (see below) were available in the ‘eyes open’ (EO) condition and for 45 TD and 38 ASD children, the data were also available in the ‘eyes closed’ (EC) condition. IQ (Mental Processing Index - MPI) has been evaluated with KABC-II (30), and the ASD participants were subdivided into two groups based on their MPI IQ scores: *average IQ* (MPI>85; ASD_>85_) and *below-average IQ* (MPI<85; ASD_<85_). Detailed information about the resulting samples is summarized in Table 1.

**Table 1.**
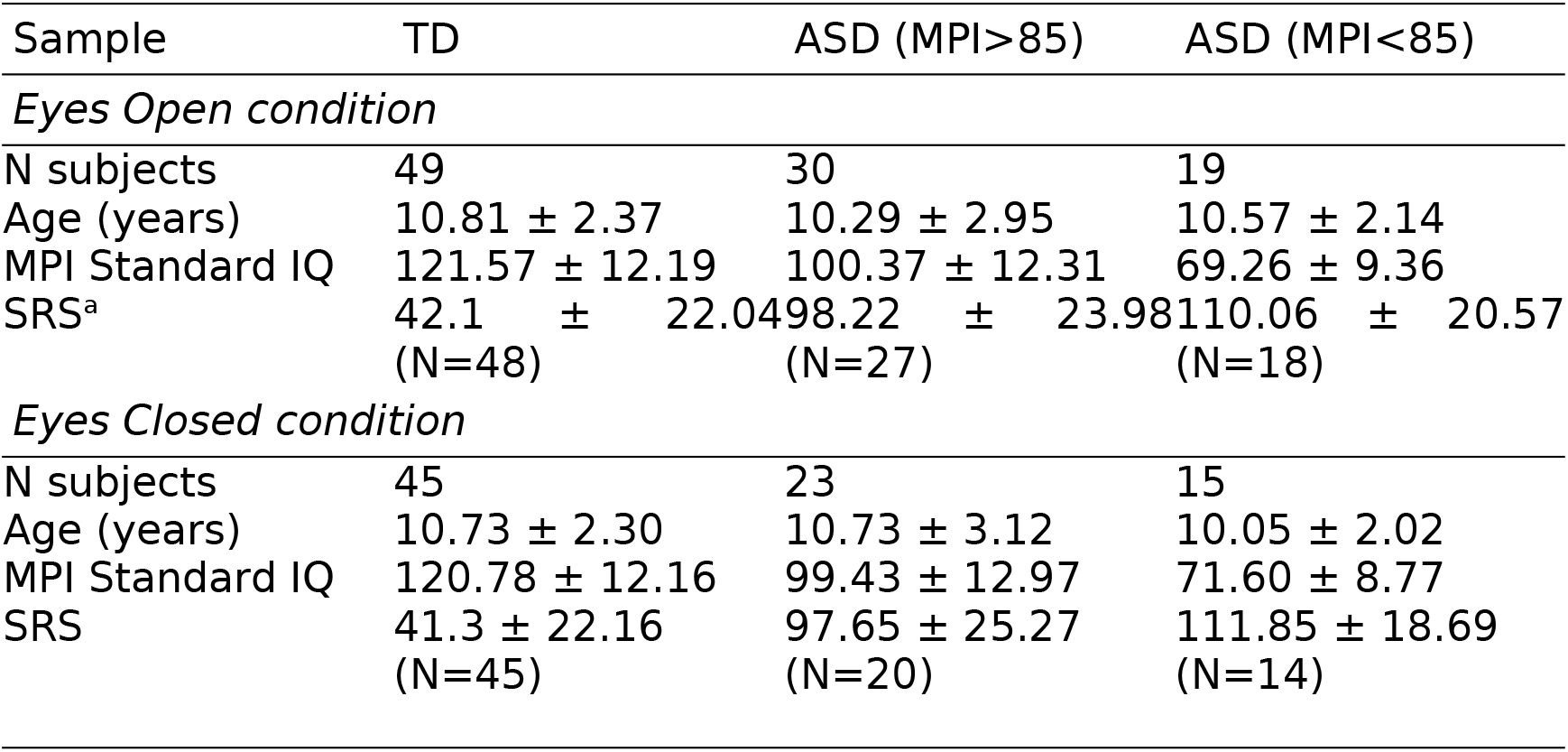
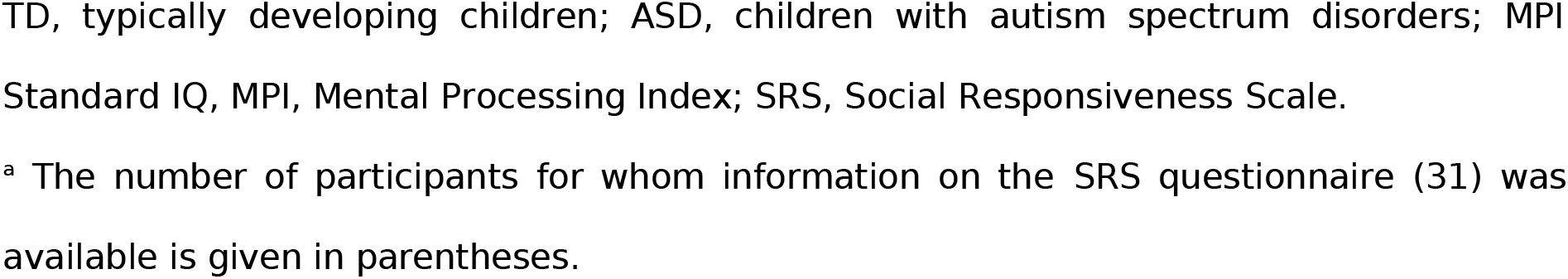
Characteristics of the experimental samples

| Sample | TD | ASD (MPI>85) | ASD (MPI<85) |
| --- | --- | --- | --- |
| *Eyes Open condition* |  |  |  |
| N subjects | 49 | 30 | 19 |
| Age (years) | 10.81 ± 2.37 | 10.29 ± 2.95 | 10.57 ± 2.14 |
| MPI Standard IQ | 121.57 ± 12.19 | 100.37 ± 12.31 | 69.26 ± 9.36 |
| SRS<sup>a</sup> | 42.1 ± 22.04<br>(N=48) | 98.22 ± 23.98<br>(N=27) | 110.06 ± 20.57<br>(N=18) |
| *Eyes Closed condition* |  |  |  |
| N subjects | 45 | 23 | 15 |
| Age (years) | 10.73 ± 2.30 | 10.73 ± 3.12 | 10.05 ± 2.02 |
| MPI Standard IQ | 120.78 ± 12.16 | 99.43 ± 12.97 | 71.60 ± 8.77 |
| SRS | 41.3 ± 22.16<br>(N=45) | 97.65 ± 25.27<br>(N=20) | 111.85 ± 18.69<br>(N=14) |

TD, typically developing children; ASD, children with autism spectrum disorders; MPI Standard IQ, MPI, Mental Processing Index; SRS, Social Responsiveness Scale.
<sup>a</sup> The number of participants for whom information on the SRS questionnaire (31) was available is given in parentheses.

### MRI data acquisition and processing

Structural MR scans (voxel size 1 mm x 1 mm x 1 mm) were performed on a General Electric Signa 1.5 T scanner. The T1 images were processed using the default FreeSurfer (v.6.0.0) ‘recon-all’ pipeline. For the MEG analysis, the cortical surface was parcellated into 448 similar-size labels, as described by Khan et al (32).

### MEG data acquisition and preprocessing

MEG was recorded with a 306-channel MEG system (Vectorview, Elekta-Neuromag). Recording was performed at rest with eyes open or closed (2-4 minutes in each condition). The signal was initially sampled at 1000 Hz and later down-sampled to 500 Hz. Head position was corrected to each subject’s initial head position using MaxFilter software (v. 2.2). The temporal signal-space separation (tSSS) method (33) with correlation limit 0.9 was applied to compensate for correlated environmental noises.

The Signal Space Projections (SSP) method (34) was applied to the raw data to suppress biological artifacts. Only gradiometers (N=204) were used for analysis. The raw signal was subdivided into 1 s non-overlapping epochs. Those epochs that were contaminated by artifacts or where the head origin deviated from the initial position by more than 20 mm were excluded from analysis. We analyzed 60 clean data epochs per condition and subject. The mean distance of the head origin from the initial position did not differ between the groups (Kruskal-Wallis one-way ANOVA: EO, H(2, N=98)=2.58, p=0.27, mean distance in TD/ASD_>85_/ASD_<85_ was 4.1/4.8/5.3 mm; EC, H(2, N=83)=0.58, p=0.75, mean distance in TD/ASD_>85_/ASD_<85_ was 2.5/2.8 /2.6 mm).

### Choice of the frequency range for the spectral slope estimation

Following previous studies (23, 25, 28), we sought to estimate the slope of aperiodic activity at the high-frequency part of the spectrum. To choose the optimal high-frequency range for the aperiodic slope analysis, we first estimated the power spectral density (PSD) in the sensor space. Figure 1 shows a power spectrum of a centrally located gradiometer (grand average over all participants) and the grand averaged power spectrum of the signal recorded by this gradiometer in the empty room.

**Figure 1.**
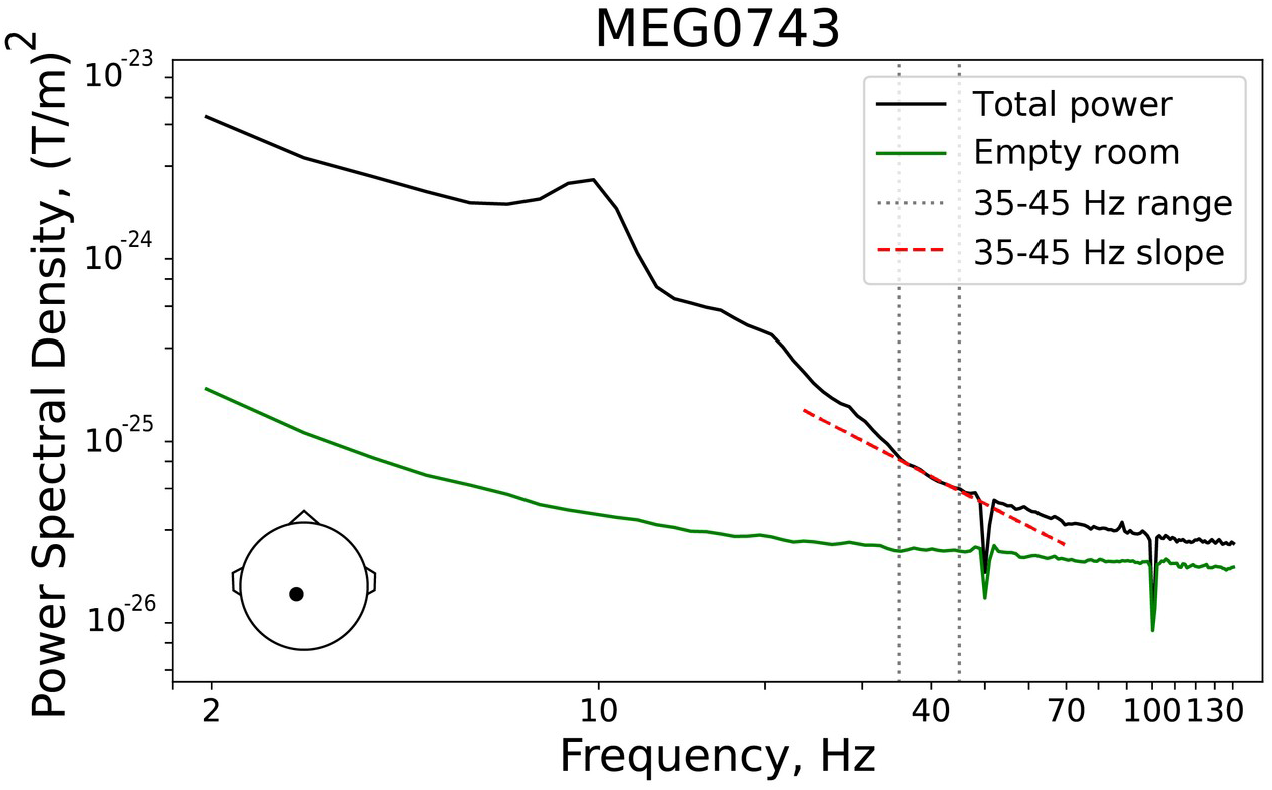
Spectral analysis for a single central gradiometer (MEG0743), positioned at a large distance from the cranial muscles: the average over all participants. Position of the gradiometer is shown in the lower left corner. Total magnetoencephalographic (MEG) power in the eyes open condition is plotted in black and the empty room noise recording is plotted in green. Spectral peaks (i.e., periodic signals) are absent in the 35-45 Hz range (marked by gray dotted lines) and the spectral power declines approximately linearly in this frequency range (red dashed line). The power spectra are plotted on a logarithmic scale.

Inspection of Figure 1 shows that the slope of the logarithmic spectrum is approximately linear between 35 and 45 Hz and deviates from the 1/f function at higher frequencies due to a decreasing signal-to-noise ratio (SNR). To minimize the contribution of periodic activity, background white noise and power line noise, we have chosen this 35-45 Hz range as the band-of-interest to estimate the spectral slope.

We performed source localization analysis using Linearly Constrained Minimum Variance (LCMV) beamformer spatial filters (35) that significantly reduce the contribution of biological artifacts (e.g., myogenic activity) into the brain activity estimated at the source level (36). We also compared the LCMV results with those obtained using standardized Low Resolution Brain Electromagnetic Tomography (sLoreta) (37) in order to gauge the effect of myogenic contamination (Supplemental Results).

### Estimation of the spectral slope in the source space

The forward model was created using a one-layer boundary element model (BEM) and surface-based source space (4096 vertices per hemisphere). The raw data were filtered between 30-140 Hz and the data covariance matrix was estimated individually for each condition. The LCMV beamformer filter with a regularization parameter of 0.1 was calculated for the source orientation that maximizes power and was applied separately to each 1 s data epoch. We then calculated the neural activity index (NAI) that represents the signal normalized to the spatially inhomogeneous noise (35). In each source vertex, the spectral power was estimated using Welch’s method. Power spectra were normalized by the corresponding maximum value and then averaged within each of the 448 labels. The spectral slope was then estimated by fitting a linear regression line to the logarithm of the power spectrum between 35 and 45 Hz vs. the logarithm of the frequency. The mean spectral slope was obtained by averaging the slopes over all cortical labels.

We also estimated the aperiodic slope using the ‘FOOOF’ parameterization algorithm (38) applied in the 2-45 Hz range. A detailed description and discussion of these results is given in the Supplemental Materials and Results.

### Estimation of the periodic power in the alpha and beta frequency ranges

Beamformers provide good estimates of spectral power changes (e.g. when contrasting conditions) (39), but might provide suboptimal estimates of the *absolute* power. Therefore, to quantify the periodic spectral power in the alpha (7-13 Hz) and beta (14-25 Hz) ranges, we performed source localization with sLoreta and then used FOOOF (38) to separate between periodic and aperiodic parts of the spectrum: periodic signal = [original spectra – aperiodic fit].

### Estimation of the sensitivity maps

To ensure that our results were not biased by head size, we calculated individual ‘sensitivity maps’ (‘mne.sensitivity_map’ function in MNE-python), which estimate how well each source is sampled by a sensor array (i.e., the planar gradiometers). The mean sensitivity in a label was then used as a nuisance variable in our regression analysis.

### Statistical analyses

To estimate the rank order stability of the spectral slopes, we calculated intraclass correlation coefficients (ICC) between conditions (eyes closed/open). ANCOVA was used to estimate group differences in the mean spectral slope as well as periodic spectral power.

Spearman correlation coefficients were calculated to estimate the link between the spectral slope and IQ. To control for nuisance variables (background noise, sensitivity, spectral power, age), we calculated partial Spearman correlations.

The False Discovery Rate (FDR) correction for multiple comparisons was applied when appropriate.

## RESULTS

### State dependency and rank order stability of the spectral slope

To become a clinically useful signature of E-I balance, the spectral slope must not only be sensitive to changes in the neural E-I ratio, but also reflect trait-like individual differences that remain stable across different functional states. In awake humans the E-I balance is shifted towards higher inhibition during states associated with high alpha power (40-42). We therefore expected that if the spectral slope in children with and without ASD is sensitive to changes in the E-I ratio, then it would be steeper (more negative) in the EC condition (characterized by stronger alpha activity), as compared with the EO condition. We also proposed that if the spectral slope represents a stable individual characteristic, it should have high rank order stability between the EO and EC conditions.

Figure 2 shows the cortical distribution of the spectral slopes in the three groups of children. First, we estimated state-related changes in the spectral slope averaged over all cortical labels (hereafter, the mean slope). A paired t-test revealed a highly significant and strong decrease of the mean slope from the EO to EC condition in the combined group of subjects (T_(82)_=5.81, p<0.000001, Cohen *d*=0.64), as well as in the separate groups (TD: T_(44)_=3.67, p=0.0006, Cohen *d*=0.55; ASD_>85_: T_(22)_=3.38, p=0.0003, Cohen *d*=0.71; ASD_<85_: T_(14)_=3.31, p=0.005, Cohen *d*=0.85; Figure 3).

**Figure 2.**
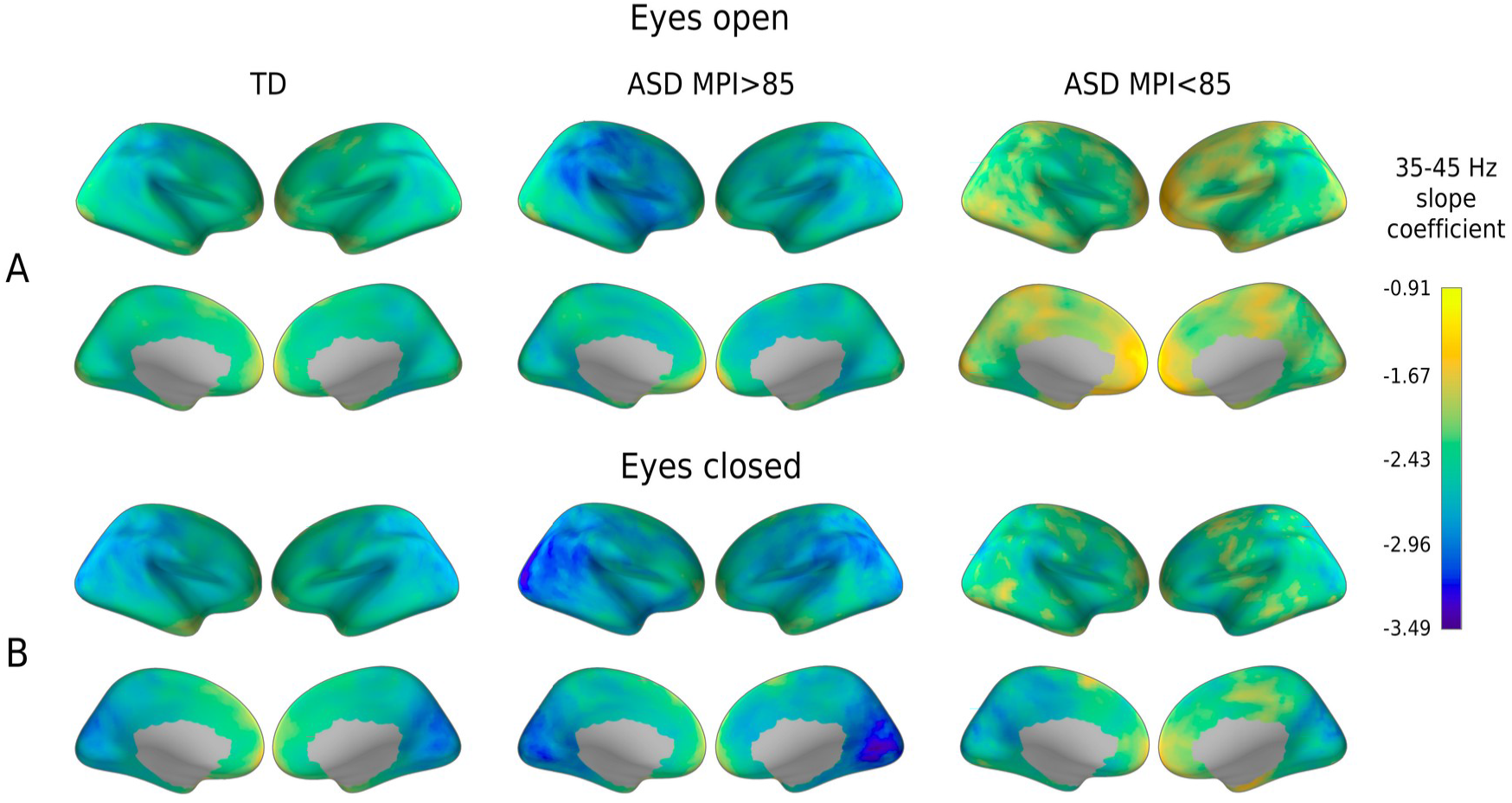
Cortical distribution of the spectral slope coefficients in the three groups of participants in the Eyes Open (A) and Eyes Closed (B) conditions. The activity of the cortical sources was estimated using the LCMV beamformer method. TD, typically developing; ASD, autism spectrum disorders; MPI, Mental Processing Index.

**Figure 3.**
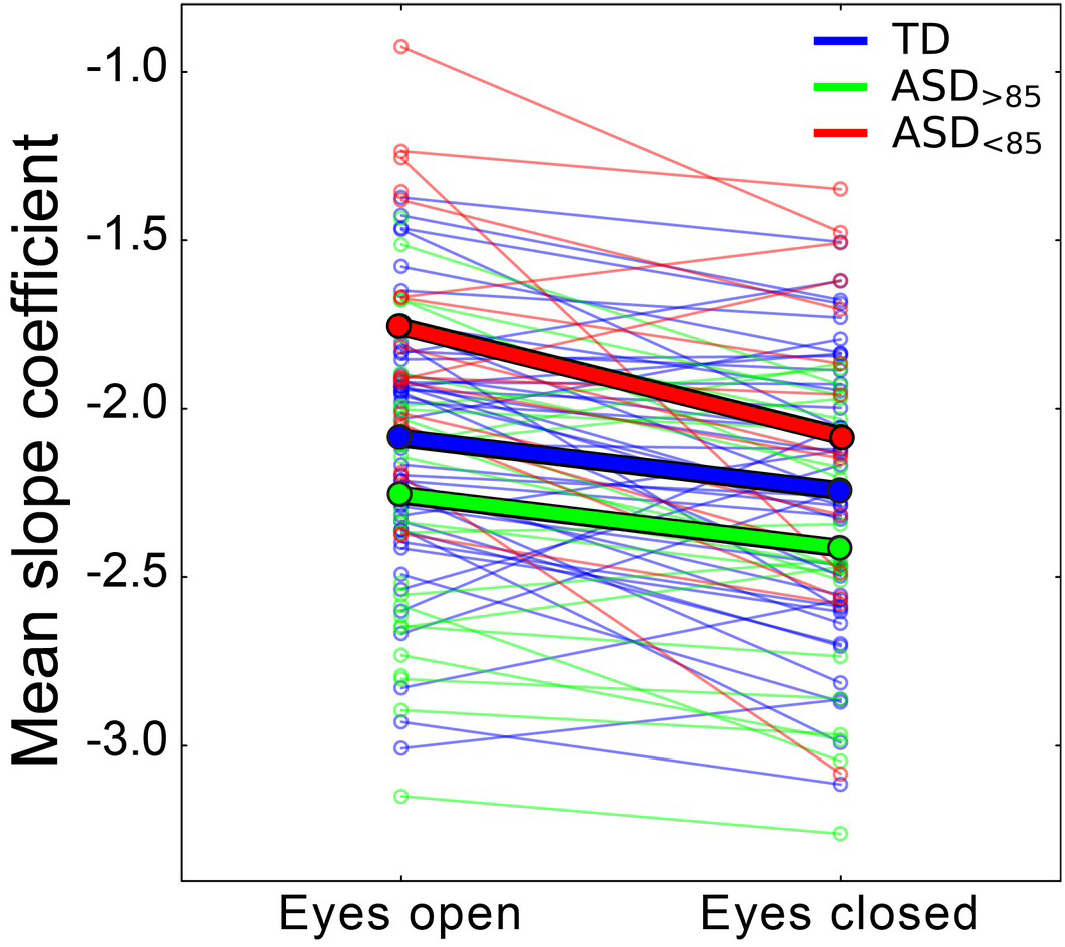
Mean spectral slope in the two experimental conditions. Thin lines show individual subjects; thick lines – the group average. Colors correspond to different groups of participants. TD, typically developing children; ASD, autism spectrum disorders; ASD_>85_, children with ASD and Mental Processing Index above 85; ASD_<85_, children with ASD and Mental Processing Index below 85.

Second, using intraclass correlations, we estimated the rank order stability of the mean spectral slope and the slopes of the separate cortical regions (448 labels) across the EC-EO conditions. The rank order stability of the mean slope was good (ICC_(82)_=0.87, p<0.000001) (43) and exceeded that in the individual cortical labels, where it ranged from 0.06 to 0.79; Figure 4). Therefore, we employed the mean spectral slope for further analysis.

**Figure 4.**
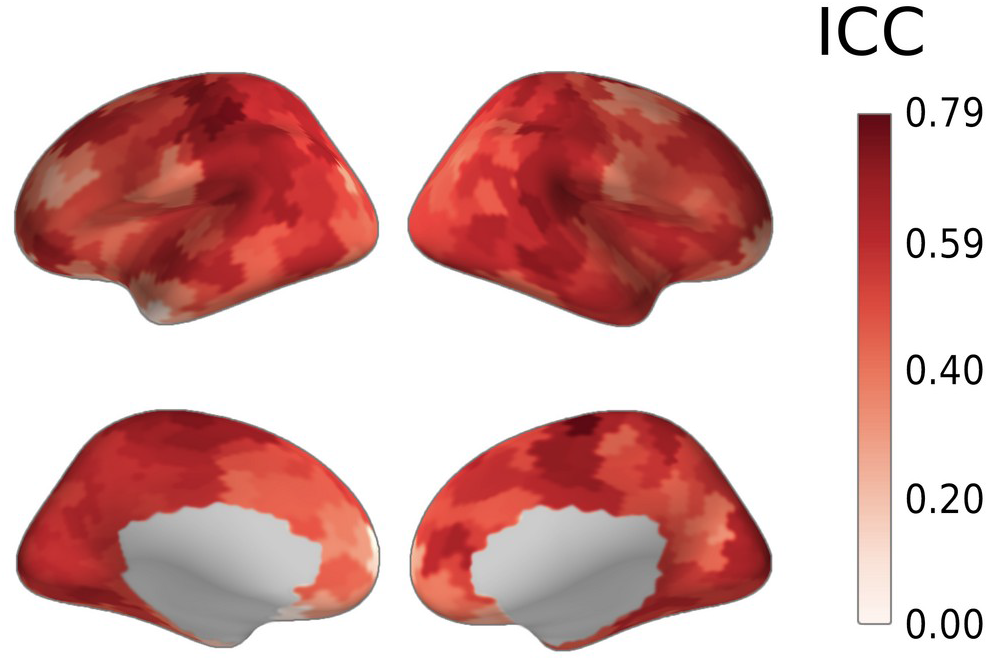
Rank order stability of the spectral slope between the Eyes Open and the Eyes Closed conditions: cortical distribution of intraclass correlation coefficients (ICC) for participants for whom data from both conditions were available (N=83).

### Effect of age, condition, IQ, and autism diagnosis on the mean spectral slope

The mean spectral slope values for the three groups of participants and the two experimental conditions are shown in Figure 5. The General Linear Model (GLM) analysis with factors Group, Condition, and Age revealed a main effect of Age (F_(1,79)_=12.76, p=0.0006, η^2^=0.14) indicating a general flattening of the spectral slope with brain maturation (Figure 6). The highly significant main effect of Condition (F_(2,79)_=38.3, p<1e-10, η^2^=0.33) did not interact with Group or Age, thus suggesting that the strong changes in spectral slope associated with opening/closing the eyes are comparable across age and in groups from different diagnostic categories. As expected, we found a significant effect of Group (F_(2,79)_=6.95, p=0.0017, η^2^=0.15): the slope was flatter in the ASD_<85_ than in either TD (F_(1,79)_=7.2, p=0.009) or ASD_>85_ (F_(1,79)_=13.8, p=0.0004) groups. Unexpectedly and by contrast, though, the slope for the ASD_>85_ group tended to be steeper than that in the TD group (F_(1,79)_=2.80, p=0.098).

**Figure 5.**
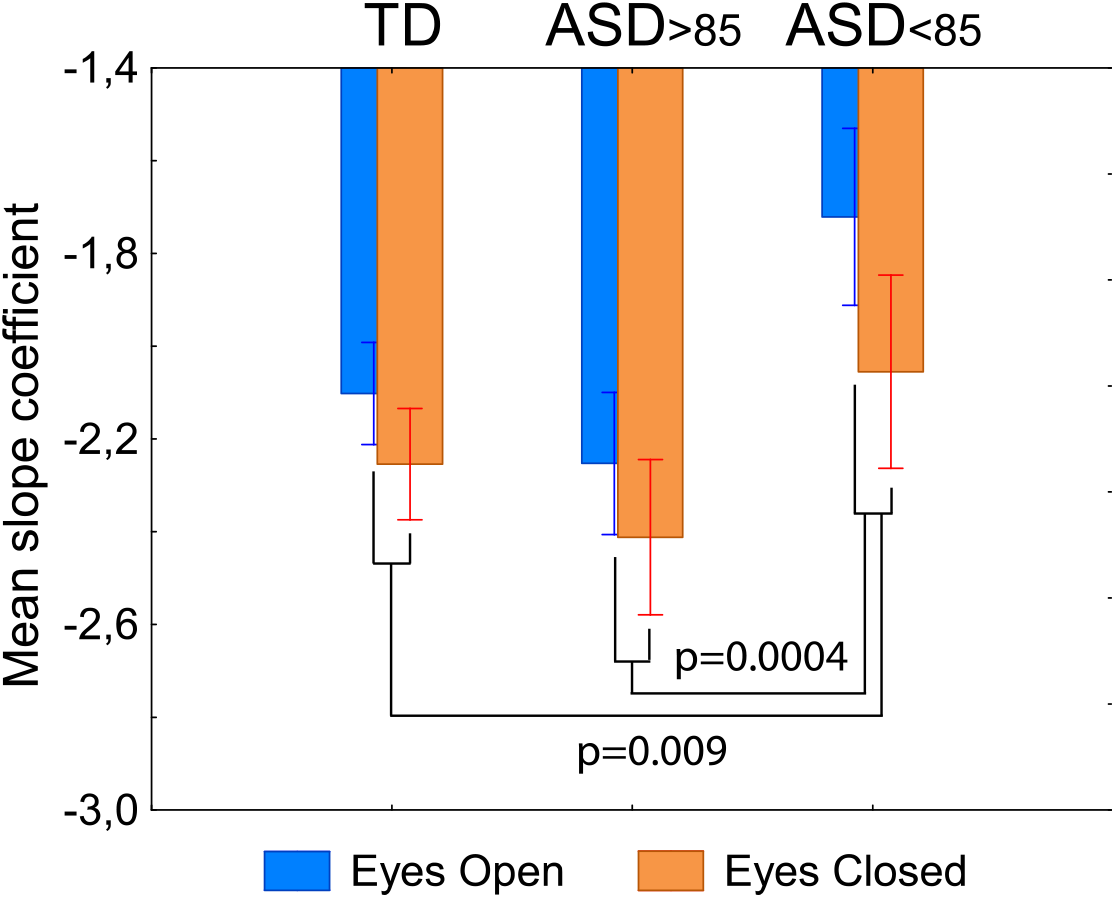
Group difference in the mean spectral slope between the three groups of participants. Vertical bars denote 0.95 confidence intervals. The p-values (two-sided t-test) are given for between group comparisons (Eyes Open and Eyes Closed conditions were pooled). TD, typically developing children; ASD, autism spectrum disorders; ASD_>85_, children with ASD and Mental Processing Index above 85; ASD_<85_, children with ASD and Mental Processing Index below 85.

**Figure 6.**
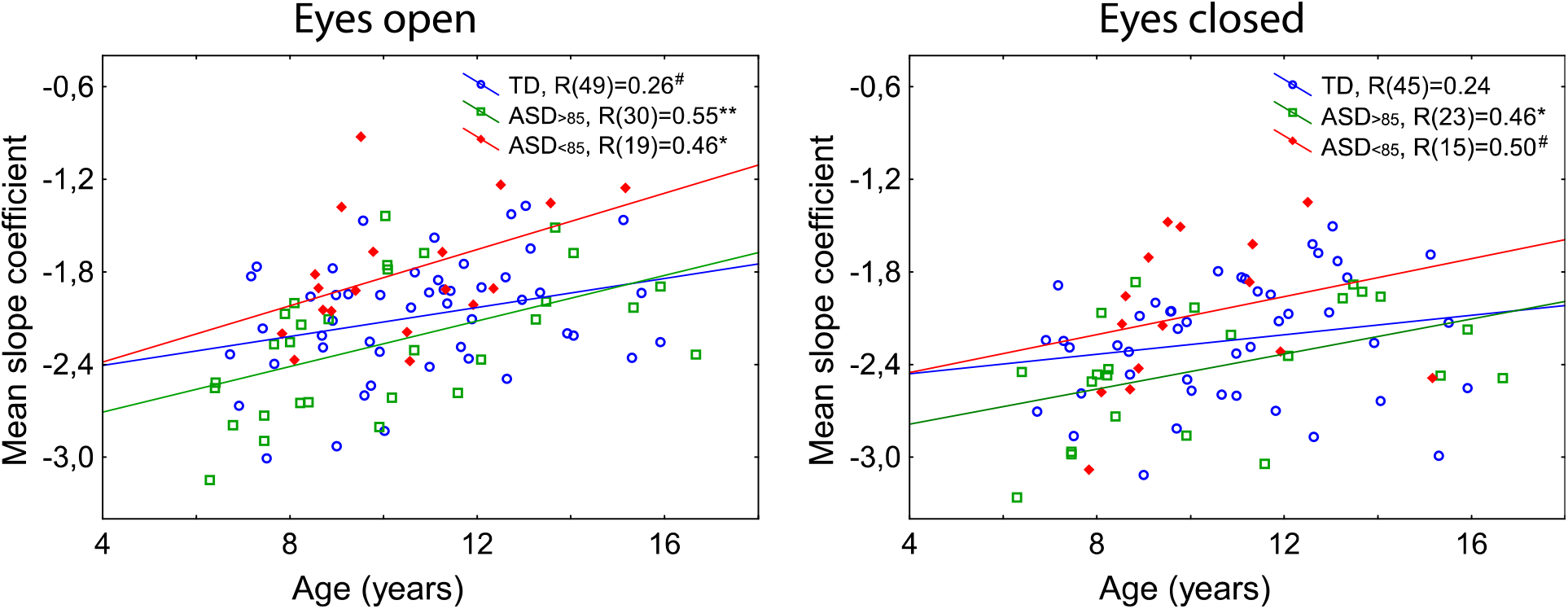
Age-related changes in the mean spectral slope in the three groups of participants. R’s denote the Spearman correlation coefficients; #p<0.1, *p<0.05, **p<0.01. The group differences between correlation coefficients (TD/ASD_<85_, TD/ASD_>85_, ASD_>85_/ASD_<85_) are not significant (all p’s>0.073). TD, typically developing children; ASD, autism spectrum disorders; ASD_>85_, children with ASD and Mental Processing Index above 85; ASD_<85_, children with ASD and Mental Processing Index below 85.

We repeated the analysis with factors Group and Age for the EO condition for which the data were available for a greater number of participants. Again, there was a significant effect of Age (F_(1,94)_=18.1, p=0.00005, η^2^=0.16) and Group (F_(2,94)_=8.4, p=0.00045, η^2^=0.15), which was explained by a flatter slope in the ASD_<85_ group than in the rest of participants.

To test if the link between the IQ and mean spectral slope present in TD children, and whether it differs from that in children with ASD, we calculated correlations between the spectral slope and IQ in the TD and combined ASD groups.

The ASD vs. TD difference in partial Spearman correlation coefficients (when adjusting for age) was significant for the EO condition (N_TD_=49, N_ASD_=49, Z=1.67, p=0.047), and the same tendency was observed for the EC condition (N_TD_=45, N_ASD_=38, Z=1.41, p=0.079). While children with ASD demonstrated reliable negative correlations between the mean slope coefficient and IQ, no such correlations were found in the TD group (Figure 7).

**Figure 7.**
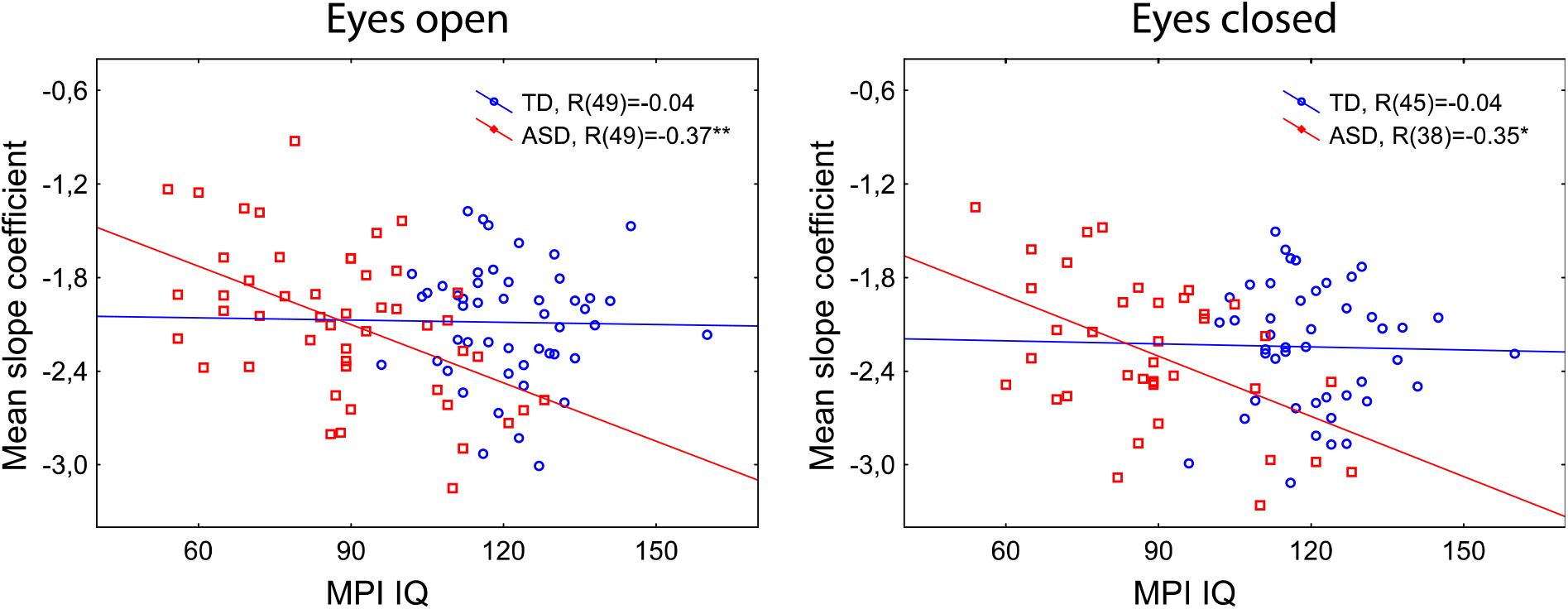
The relationship between Mental Processing Index (MPI IQ) and mean spectral slope in typically developing (TD) children and in children with autism spectrum disorders (ASD). R’s denote the partial Spearman correlation coefficients, adjusting for age. *p<0.05, **p<0.01.

### Cortical distribution of the correlations between spectral slope and IQ in children with ASD

To ensure that the link between the mean spectral slope and IQ is not driven by brain regions wherein source estimates are most susceptible to myogenic contamination, we calculated partial Spearman correlation coefficients between MPI IQ and spectral slopes in the separate cortical labels, while controlling for age and sensitivity (see Methods for details). For the EO condition, the largest group of significant correlations (p<0.05, FDR corrected) overlapped the supramarginal area and temporoparietal junction of the right hemisphere (Figure 8, see Supplemental Table S2 for the full list of the cortical labels that survived the FDR correction). In general, the spatial distribution of the correlations is evidence against a potential ‘myogenic explanation’ of their origin. Similar negative correlations were observed for the EC condition, but none of them survived correction for multiple comparisons (Figure 8).

**Figure 8.**
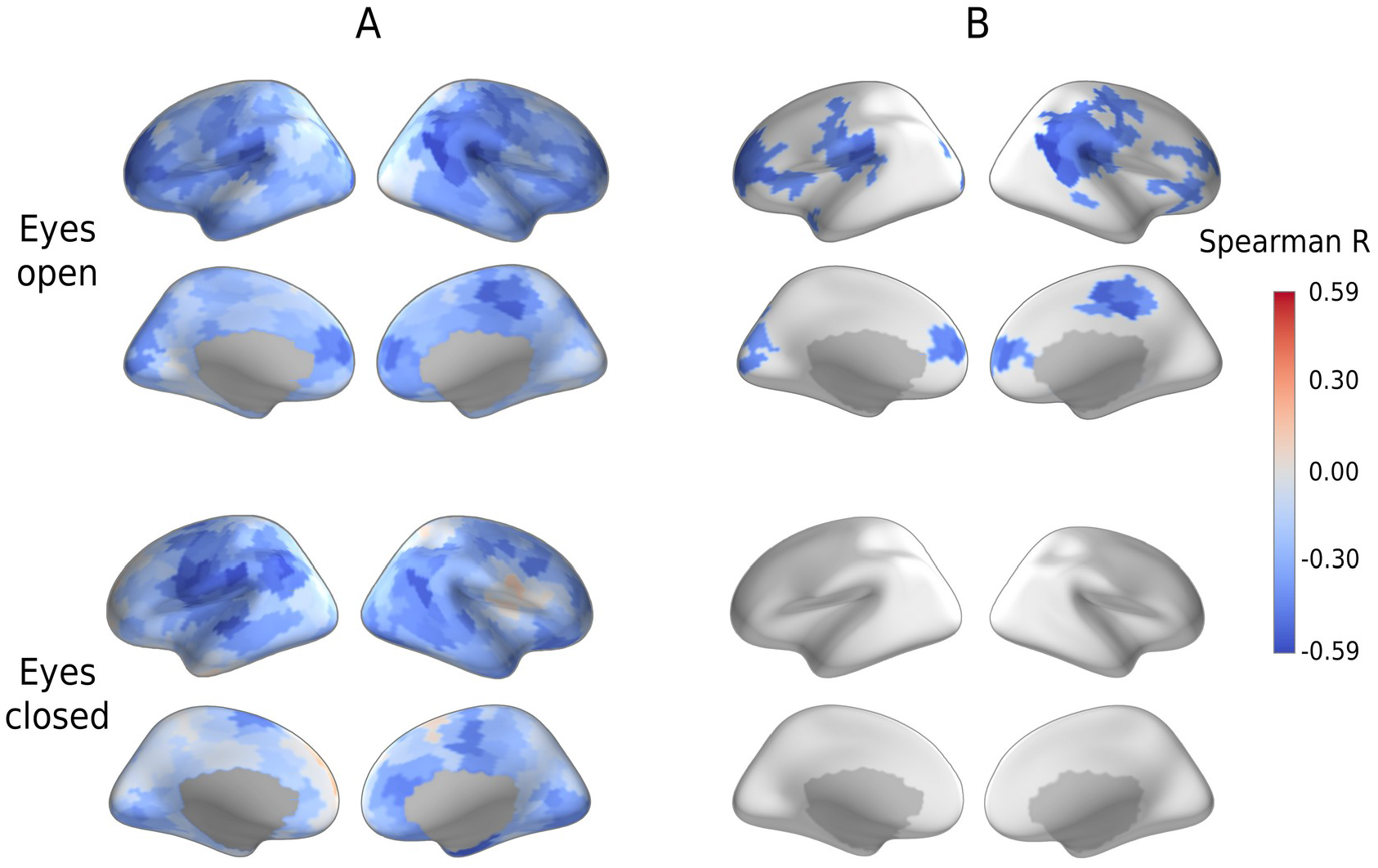
Partial Spearman correlations between spectral slopes in the individual cortical labels and Mental Processing Index (MPI IQ) in children with autism spectrum disorders (ASD) in the Eyes Open and Eyes Closed conditions, adjusting for age and label-specific sensitivity. (A) Uncorrected for multiple comparisons. (B) False Discovery Rate corrected.

### Spectral slope and periodic activity in the alpha and beta range

It has been argued that changes in the spectral slope of brain activity are secondary to the changes in periodic activity (e.g. power of alpha or beta rhythms) (44). The power of periodic activity varies with functional and behavioral state, which in turn may depend on a subject’s diagnosis and IQ. To ensure that the group differences in the spectral slope were not explained by differences in the periodic activity, we estimated the periodic spectral power in the alpha and beta bands.

Figure 9 shows cortical distributions of alpha and beta periodic power in the three groups of participants. In accord with the results of Muthukumaraswamy and colleagues (44), a stronger periodic alpha and beta power was associated with a steeper, more negative spectral slope in both TD and ASD participants and in both experimental conditions (Table 2). However, unlike the slope, the grand averaged periodic power did not differ between the groups (ANOVA Group effect for the alpha band, EO: F_(2,94)_=1.89, p=0.16, η^2^=0.038; EC: F_(2,80)_=1.29, p=0.28, η^2^=0.031; ANOVA Group effect for the beta band, EO: F_(2,94)_=1.4, p=0.26, η^2^=0.028; EC: F_(2,80)_=2.1, p=0.13, η^2^=0.049).

**Table 2.**
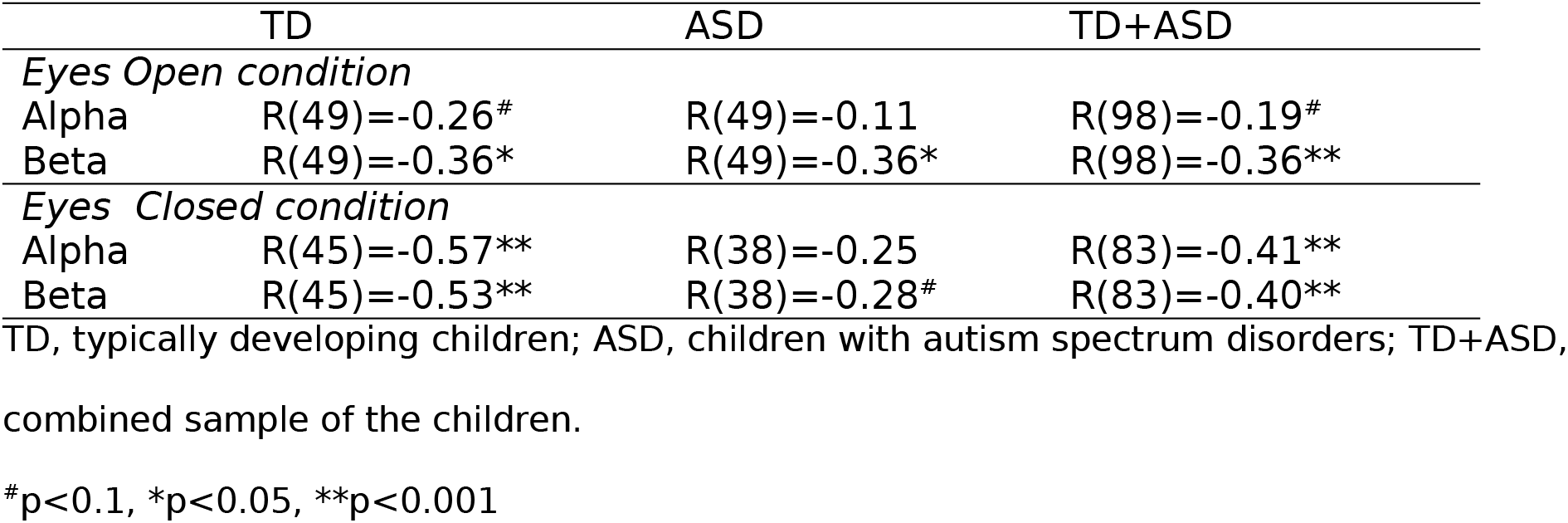
Spearman correlations between the grand average spectral slope and power of periodic oscillations in the alpha (7-13 Hz) and beta (14-25 Hz) bands

**Figure 9.**
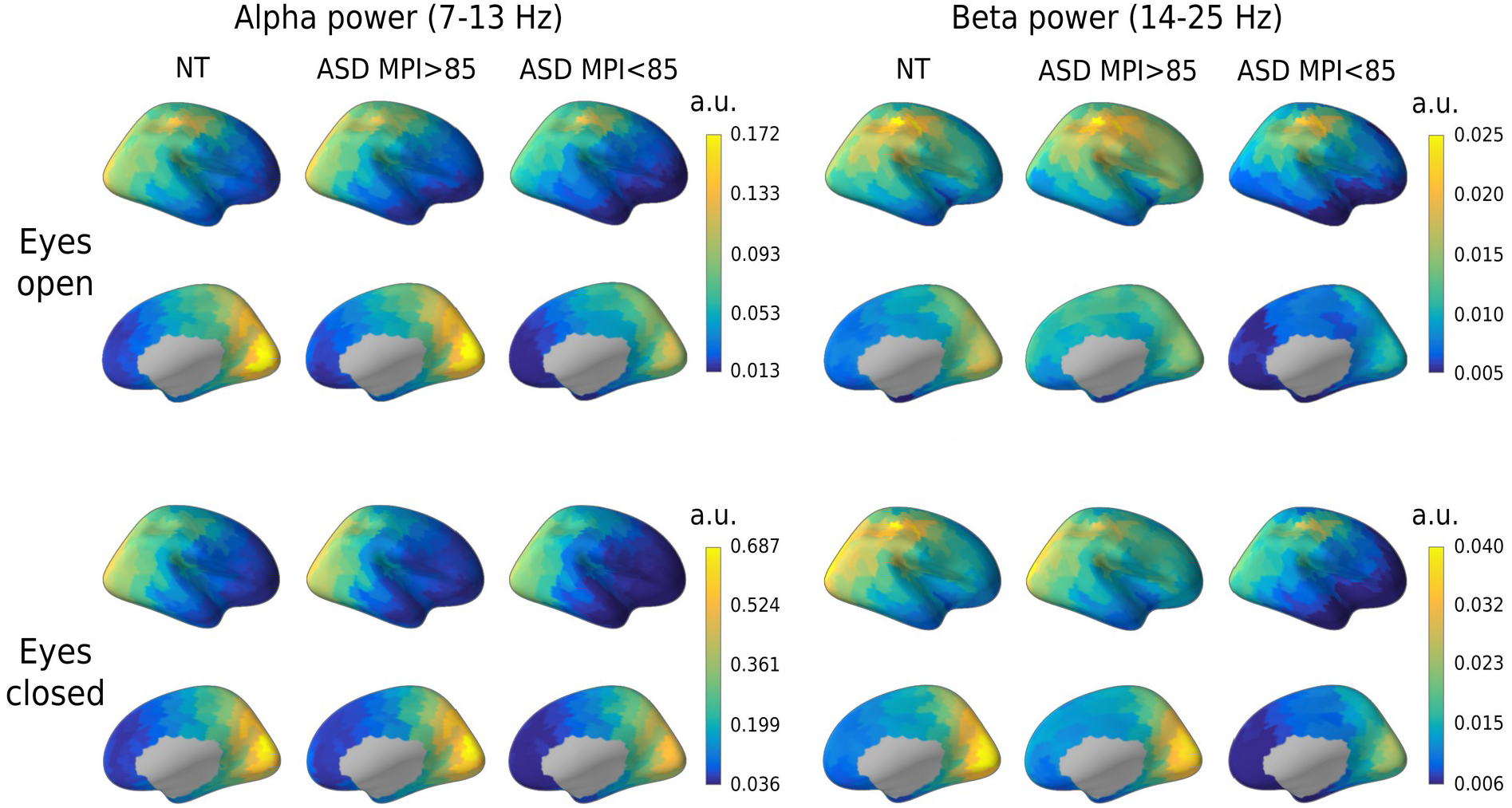
Cortical distributions of the power of periodic activity in the alpha and beta bands in the three groups of participants and in the Eyes Open and Eyes Closed conditions. Note that the scales differ for the bands and conditions. TD, typically developing children; ASD, autism spectrum disorders; ASD_>85_, children with ASD and Mental Processing Index above 85; ASD_<85_, children with ASD and Mental Processing Index below 85.

To further ensure that the link between spectral slope and IQ in the ASD group did not stem from inter-individual differences in periodic activity, we calculated partial Spearman correlations between the mean spectral slope and IQ while controlling for the mean alpha and beta power. Partialing out the periodic power did not significantly affect the correlation between the mean spectral slope and MPI IQ (EO: N=49, R_partial_=-0.43, p=0.003; EC: N=38, R_partial_=-0.35, p=0.04).

## DISCUSSION

The power of electrophysiological brain activity exhibits a 1/f-like dependence, and the flattened slope of the power decay as a function of frequency has been previously linked to a shift in the neural E-I balance towards excitation (22-26). Here, we found that the spectral slope (averaged over cortical regions) was flatter in children with ASD and below-average IQ than in either TD children or in those with ASD and average IQ (Figure 5). This finding suggests that the neuronal E-I balance in the brains of the below-average IQ children with non-syndromic ASD is shifted toward excitation at the global scale.

Since the balanced activity of excitatory and inhibitory neurons is necessary for normal development and functioning of the brain (45-47), changes of the E-I ratio may contribute to intellectual disability in ASD. Indeed, mutations in the genes important for neural communication and the E-I balance may lead to both autism and intellectual disability (11-13, 48, 49). Moreover, the putative shift to neuronal hyper-excitability observed in children with below-average IQ in our study may explain the high prevalence of epilepsy at the low-IQ end of the autism spectrum (14, 15).

The correlation of the spectral slope with IQ characterized children with ASD, but not TD children (Figure 7). It is noteworthy that in the ASD group these correlations formed a large right-hemispheric cluster (Figure 8), which included the temporoparietal junction and supramarginal gyrus – areas previously implicated in the theory of mind (50, 51) and empathy (52), respectively. Functional abnormalities in these areas were frequently found in ASD in fMRI studies (53-56). Moreover, the whole set of regions where the flattened spectral slope is linked to poor performance IQ (Supplemental Table S2) essentially overlaps with the frontoparietal executive network, whose abnormality is critically involved in executive dysfunction and the profound decrease in adaptive functioning in ASD (for a meta-analysis of fMRI findings see (57)). The spatial information in the present study, however, should be interpreted with caution given the unequal sensitivity of the MEG sensors to the activity of different cortical sources and the relatively low intraclass correlations of the region-specific spectral slopes between experimental conditions (Figure 4).

No significant differences in the spectral slope were found between children with ASD and average IQ and those with typical development. Interestingly, children with ASD and IQ above 85 display a tendency to have a steeper mean spectral slope than the TD children. It is likely that in the less severely affected children with ASD, the E-I imbalances are compensated on the global scale (or even biased towards inhibition) due to homeostatic plasticity mechanisms (7).

Although the mean spectral slope reliably differentiated children with ASD and intellectual disability from the rest of the sample, these children share some properties of 1/f slope that are also pertinent to both average-IQ children with ASD and their TD peers.

Firstly, the mean spectral slope becomes less negative with age in all groups of participants (Figure 6). The developmental flattening of the spectral slope has been previously observed in TD infants (21) and children (19, 20) and may reflect protracted maturational changes that are common for both TD and ASD individuals. These maturational changes do not necessarily reflect changes in neural E-I balance, since the 1/f slope is sensitive to a multitude of processes (17), that include not only developmental changes in the number and composition of GABA and glutamatergic receptors (58, 59), but also axon myelination (60) and changes in cortical thickness (61). Considering the strong age dependence of the spectral slope, age should be carefully controlled for in studies employing the slope as a potential biomarker of the E-I balance.

Secondly, in both TD and ASD the spectral slope was steeper in case of stronger alpha and beta oscillations (Table 2) and generally steeper during the EC condition associated with high power in the alpha rhythm (Figure 3). These findings are in line with a previous report (44) and may stem from a partial overlap in the inhibitory mechanisms affecting alpha oscillations and the spectral slope. Indeed, ‘alpha states’ index reduced cortical excitability (40, 41) and are dominated by currents and firing in supragranular cortical layers (62), which display steeper 1/f slope than the other cortical layers, presumably due to longer time constants and a higher density of inhibitory transmembrane currents (63). Therefore, inhibitory currents in the supragranular layers may both promote alpha oscillations and enhance the 1/f power decay. However, partialling out the variability in periodic power did not change the negative correlation of the spectral slope with IQ in ASD. This finding clearly indicates that the spectral slope brings unique information that is different from that which can be derived exclusively from the periodic activity.

Importantly, the correlations of the spectral slope with IQ in our study are unlikely to be explained by differences in biological or instrumental noise (see Supplemental Results for an analysis of the contribution of instrumental noise and muscle artifacts to the spectral slope and power in the 35-45 Hz range). Therefore, the link between IQ and the slope of the aperiodic component of the MEG-detected neural activity spectrum in children with ASD seems to be a true finding that reflects a globally elevated neuronal excitability associated with intellectual disability that often co-occurs with autism.

There are, however, a few potential improvements that could increase the reliability of this measure and promote its practical application. *Firstly*, while our participants were not engaged in any specific activity, one should consider using a modified experimental paradigm that allows for longer periods of artifact free data in children (64). *Secondly*, the accuracy of the slope estimation could be improved by increasing the upper limit of the frequency range beyond 45 Hz, e.g. when the power line cycle is 60 Hz and/or a better SNR is available. *Thirdly*, the new and rapidly developing technology of ‘on-scalp’ MEG (65-67) may help to obtain equally high-quality data in all parts of the head, including the frontal areas, which are typically positioned furthest away from the sensors in ‘conventional’ MEG systems.

An important limitation of our study is the high heterogeneity of our ‘idiopathic’ ASD sample that included subjects with different etiologies of ASD. To confirm the validity of the spectral slope biomarker, it would be important to investigate participants with monogenic disorders and animal models that are characterized by predominant shifts of the E-I balance towards excitation (8-10) or inhibition (5, 6). Another caveat is that putative abnormalities of the E-I ratio inferred here by the 1/f slope may not be specific to autism, but rather play a general role in the pathophysiology of intellectual disability. Future research should explore this question.

In conclusion, the abnormally flattened 1/f spectral slope estimated in the high-frequency part of the MEG-detected neural activity spectrum is likely to reflect neuronal E-I imbalance associated with intellectual disability in children with ASD. Participants with below-average IQ are heavily underrepresented in autism research (68) and most neuroimaging studies only include high-functioning individuals with ASD (16). Our MEG study demonstrates that it is feasible to collect resting-state MEG and structural MRI data in children with below-average IQ, and that cognitive ability, even though it is not a core aspect of the ASD diagnosis per se, should be considered as an important factor for research in the pathophysiology of this neurodevelopmental disorder. Our results strongly indicate that the 1/f spectral slope estimated in the high-frequency part of the neural activity power spectrum may be a useful and objective biomarker of changes to the E-I ratio induced by pharmacological and other therapeutic interventions in low-functioning children with ASD.

## Data Availability

All data produced in the present study are available upon reasonable request to the authors

## Acknowledgements

A special thanks to all of the families and participants who join with us in this effort. The study has been supported by the Moscow State University of Psychology and Education (MSUPE).

