## Supplementary Information for "Globally elevated excitation-inhibition ratio in children with autism spectrum disorder and below-average intelligence"

### SUPPLEMENTARY METHODS AND MATERIALS

(full version of **METHODS AND MATERIALS**)

#### Participants

Fifty-eight typically developing (TD) boys and 63 boys with autism spectrum disorders (ASD), aged 6 to 15 years, were initially enrolled in this study. The TD children were recruited from local schools in Moscow and had no neurological or psychiatric disorders. The ASD children were recruited from rehabilitation centers affiliated with the Moscow University of Psychology and Education. Each participant in the ASD group had a diagnosis of ASD confirmed by an experienced psychiatrist according to the DSM-5 criteria and interviews with children's parents or caregivers. Parents/caregivers of all children were asked to fill in the Russian version of the Social Responsiveness Scale for children (SRS) (1). Intelligence quotient (IQ) has been evaluated through standard scores on K-ABC subscales (Simultaneous and Sequential), as well as by calculating the Mental Processing Index (MPI) (2). The study has been approved by the Ethical Committee of the Moscow State University of Psychology and Education. Verbal assent to participate in the experiment was obtained from all the subjects; parents/caregivers provided informed written consent.

After visual inspection of MEG recordings, part of the children (9 and 13 TD, 14 and 25 ASD in the 'eyes open' (EO) and 'eyes closed' (EC) conditions, respectively) were excluded from further MEG data analysis due to excessive muscle or other artifacts, or due to too large displacement of the head origin from the initial position (see below). The final number of participant datasets used for analysis was 49 TD and 49 ASD in the EO condition and 45 TD and 38 ASD in the EC condition. The TD and ASD participants did not differ in age

(Mann-Whitney U test, EO condition: $N_{TD}$=49, $N_{ASD}$=49, U=1014.5, Z=1.31, p=0.19; EC condition: $N_{TD}$=45, $N_{ASD}$=38, U=745.0, Z=1.15, p=0.25), but differed in the MPI scores (Mann-Whitney U test, $p<10^{-10}$ for both conditions). The SRS scores were significantly higher in participants with ASD than in the control sample (Mann-Whitney U test, $p<10^{-10}$ for both conditions).

For further analysis, ASD participants were subdivided into two groups based on the MPI scores: *average IQ* (MPI>85; $ASD_{>85}$) and *below-average IQ* (MPI<85; $ASD_{<85}$). Detailed information about the resulting samples is summarized in Table 1 in the main manuscript. The SRS scores tended to be higher in children with ASD and below-average IQ ($ASD_{<85}$) than in those with ASD and average IQ ($ASD_{<85}$) (T-test, EO: $N_{ASD>85}$=27, $N_{ASD<85}$=18, $t_{(43)}$=1.71, p=0.09; EC: $N_{ASD>85}$=20, $N_{ASD<85}$=14, $t_{(32)}$=1.78, p=0.08), but age did not differ in the two ASD groups (Mann-Whitney U test, p's>0.3 for both conditions).

#### MRI data acquisition and processing

Structural MR scans (voxel size 1 mm x 1 mm x 1 mm) were performed on a General Electric Signa 1.5 T scanner. The obtained T1 images were processed using the default FreeSurfer (v.6.0.0) 'recon-all' pipeline. For the MEG analysis, the cortical surface was parcellated into 448 similar-size labels, as described by Khan et al (3).

#### MEG data acquisition and preprocessing

MEG was recorded at the MEG Center of the Moscow State University of Psychology and Education with a 306-channel MEG system (Vectorview, Elekta-Neuromag). The 'resting state' recordings were always performed in the beginning of the recording session followed by various tasks. The participants were instructed to sit calmly, first with eyes closed and then they were instructed to open their eyes. No other instructions were given. 2-4 minutes of MEG were recorded in each experimental condition. Participants' behavior was constantly monitored through a video camera positioned inside the MEG room. The operator started the MEG recording as soon as the subject fulfilled the instruction to sit calmly. Trigger marks and experimenter's comments were further used to select MEG intervals for analysis.

The signal was initially sampled at 1000 Hz with a 0.03-300 Hz band-pass filter. Head position was continuously monitored and the position information was later used to compensate for potential head motion by converting the signal to each subject's initial head position using MaxFilter software (v. 2.2). The temporal signal-space separation (tSSS) method (4) with correlation limit 0.9 was applied to compensate for correlated environmental noises.

Further steps of data preprocessing were performed with the MNE-python toolbox (v.0.22) (5). The raw data were down-sampled to 500 Hz. If several segments of the data were available for the same condition, they were joined into one 'raw' file and the Signal Space Projections (SSP) method (6) was applied to the raw data to suppress biological artifacts. One electrooculogram and one cardiac projection were excluded for each subject and condition (EO, EC). The raw signal was then subdivided into 1 s non-overlapping epochs. Epochs contaminated with bursts of myogenic artifacts were automatically detected using MNE-python function 'annotate_muscle_zscore' with default parameters (ch_type='mag', threshold=5.0, min_length_good=0.2, filter_freq=[110, 140]) and excluded from analysis. From this point on, only gradiometers were used for analysis. All epochs where then visually inspected and those containing obvious artifacts (jumps in the data, bursts of myogenic artifacts undetected automatically) were excluded. We also excluded those epochs where the head origin deviated from the initial position by more than 20 mm. The percent of dropped epochs was higher in the ASD than in TD children in the EO condition (Mann-Whitney U=940.0, p=0.03) and tended to be higher in the EC condition (Mann-Whitney U=733.0, p=0.10). To preclude group differences in timing of the analyzed epochs relative to the condition onset, we randomly excluded 10% of the 'good' epochs in the TD group and then have chosen the first 60 epochs per condition and subject in both ASD and TD groups for further analysis. As a result, there was no significant difference in epoch times relative to the condition onset time between the three experimental groups for both EO (Kruskal-Wallis one-way ANOVA, H(2, N=98)=2.73 p=0.25) or EC (Kruskal-Wallis one-way ANOVA, H(2, N=83)=2.38, p=0.30) conditions. The mean distance of the head origin from its initial position during MEG recording did not differ between the groups (Kruskal-Wallis one-way ANOVA: EO, H(2, N=98)=2.58, p=0.27, mean distance in TD/$ASD_{>85}$/$ASD_{<85}$ was 4.1/4.8/5.3 mm; EC, H(2, N=83)=0.58, p=0.75, mean distance in TD/$ASD_{>85}$/$ASD_{<85}$ was 2.5/2.8 /2.6 mm).

**Empty room recording**

To estimate background noise, MEG data was recorded in the absence of a subject (empty room), directly before each subject's MEG recording. The empty room MEG recordings were spatially filtered using the tSSS method (4) and were further used to estimate the background noise in the frequency range of interest, as well as to construct the noise covariance for the sLoreta analysis (7), as described below.

**Choice of the frequency range for the spectral slope estimation**

Following previous studies (8, 9), and in order to avoid complications associated with delta peak that may present in some participants at the lower edge of the analyzed frequency range (10), we sought to estimate the slope of aperiodic activity at the high-frequency part of the spectrum. To that end, we inspected the power spectral density (PSD) in sensor space (Welch's method; see below). Figure 1 in the main manuscript shows power spectra of a centrally located gradiometer for the EO (Total power) and empty room (noise) datasets (grand averaged over all participants). Importantly, we selected a centrally-located gradiometer for this analysis because of its relatively large distance from cranial muscles, meaning it is relatively insensitive to myogenic artifact sources.

The overall 'total power' spectrum presents a typical 1/f-like characteristic with peaks corresponding to the typical neuro-oscillation bands. The effect of these periodic signals is most significant at low frequencies (e.g., the alpha at approximately 10 Hz); for frequencies above approximately 35 Hz, peaks are not obvious and the 1/f-like trend is apparent. We therefore set the lower bound for our aperiodic activity slope estimate to 35 Hz. On the other hand, a knee appears in the spectrum for frequencies above approximately 50 Hz where the 'total power' approaches that of the empty room spectrum (i.e., where power is approximately constant as a function of frequency). In order to also avoid potential effects of the 50 Hz power line cycle and the notch filter used for its removal, we set the upper bound for our aperiodic activity slope estimate to 45 Hz. In the 35-45 Hz range, the grand averaged over all subjects in EO condition spectrum displays a clear 1/f-like characteristic (i.e., in the 35-45 Hz range, the 'total power' spectrum is well-approximated by a line on the log-log scale of Figure 1 in the main manuscript).

Yet another potential source of noise that may confound the spectral slope estimation in the high-frequency range is the activity of cranial muscles.

To decrease the influence of myogenic artifacts, we performed source localization analysis using Linearly Constrained Minimum Variance (LCMV) beamformer spatial filters that significantly reduce the contribution of muscle artifacts in the brain activity estimated at the source level (11). In order to gauge the effect of myogenic contamination on the spectral slope in the 35-45 Hz frequency range, we also compared the results obtained using two source localization methods that are understood to provide different suppression factors for myogenic contamination (LCMV beamformer vs. Standardized Low Resolution brain Electric Tomography (sLoreta); see section 'Putative contribution of myogenic artifacts to the 35-45 Hz spectral slope and its group differences' below).

**Estimation of the spectral slope**

We performed co-registration of the MEG data to the magnetic resonance (MR) images using the mne_analyze tool (MNE-C software; (12)). The forward model was created using a one-layer boundary element model (BEM) and surface-based source space (4096 vertices in each hemisphere). The raw data were filtered between 30-140 Hz, and the same epochs as those selected at the preprocessing step were used for the source analysis.

While the main analysis was conducted using LCMV beamformer (13), to estimate the effect of the source localization method we also repeated the analysis using sLoreta (7).

For LCMV beamformer, the data covariance matrix was estimated individually for each condition using method 'shrunk'. No noise covariance based spatial whitening was applied. The resulting filter with a regularization parameter of 0.1 was calculated for the orientation that maximizes power and was applied separately to each 1 s data epoch. We then calculated the neural activity index (NAI) that represents the signal normalized to the spatially inhomogeneous noise (13).

For the sLoreta analysis, the noise covariance matrix was calculated from one minute of ‘MaxFiltered’ and band-passed (30-140 Hz) empty room data. The sLoreta inverse solution was estimated with the following parameters: semi-orthogonal orientation of the dipole source to the cortical surface (parameter loose set to 0.4), depth weighting parameter set to 0.8, and signal-to-noise ratio (SNR) set to 1.

To estimate spectral power at the source level, the Welch’s method was applied to the resulting source estimates (‘virtual sensors’). The data epochs were smoothed with a Hamming taper, and a Fast Fourier Transform with a frequency resolution 1 Hz was performed for the time series at each vertex (‘virtual sensor’) to estimate the PSD across the epochs.

The power spectra were averaged within each of 448 labels (see above). In order for the spectra of each source vertex to contribute equally to the average, the power spectra in each vertex were normalized by the corresponding maximum value before averaging. For each label, we calculated the spectral slope by fitting a linear regression line to the logarithm of the power spectrum between 35 and 45 Hz vs. the logarithm of the frequency. For this, we used the ‘polyfit’ function from the Python library ‘NumPy’ (v.1.20.2). The linear term coefficient was used as an estimate of the 35-45 Hz spectral slope (hereinafter referred to as the ‘spectral slope’).

Recently, the power spectrum parameterization algorithm (Fitting Oscillations One Over F – FOOOF) has been proposed, that allows separation of the periodic and aperiodic components of the spectrum in a broad frequency range (14). Since this algorithm is increasingly used in the M/EEG field, we compared the results obtained with the FOOOF algorithm applied in the broad frequency range (2-45 Hz) to those obtained by direct estimation of the spectral slope in the high-frequency range (35-45 Hz). The detailed description of the method and results for this broadband data analysis are given in the separate section below ('Spectral slope estimated with FOOOF').

**Estimation of the periodic power in the alpha and beta frequency ranges**

The normalized beamformer solution (NAI) is not always effective for estimating absolute power (see an example at 'https://natmeg.se/ft_beamformer/beamformer.html'). Therefore, for the source estimation of the absolute alpha (7-13 Hz) and beta (14-25 Hz) powers, we have chosen to use sLoreta, which has zero localization error (7). Unlike the beamformer approach, source localization with sLoreta does not suppress biological artifacts. However, since the MEG signal in the alpha and beta bands is not strongly contaminated by myogenic artifacts (15), the method's capacity to suppress these artifacts is not critically important in the case of these low-frequency rhythms.

For the low-frequency power analysis, the signal was band-passed in the 1-47 Hz range. The noise covariance matrix was derived from one minute of 'MaxFiltered' and band-passed (1-47 Hz) empty room data. The sLoreta inverse solution was estimated with the following parameters: semi-orthogonal orientation of the dipole source to the cortical surface (parameter loose set to 0.4), depth weighting parameter set to 0.8, and SNR set to 1. The inverse operator was applied separately to each one-second data epoch and the spectral analysis was performed at the source level as described above.

To separate the periodic from aperiodic spectral power in the alpha and beta bands, we applied the FOOOF algorithm to the average power spectra of each label in the 2-45 Hz range. Even if this method may not provide a perfect estimation of the aperiodic slope in our study (see section 'Spectral slope estimated with FOOOF' below), it gives a reasonable approximation on the power of periodic component in alpha and beta ranges, as one can see from the Figure 9 in the main manuscript. The periodic component was then estimated as: [original spectra – aperiodic fit]. The mean alpha and beta powers were assessed as an average spectral power of the periodic component in the 7-13 Hz and 14-25 Hz bands, respectively.

##### Estimation of the sensitivity maps

The sensors' sensitivity to the brain sources is inversely proportional to the square of the distance from the source to the sensor ($1/\text{distance}^2$), and is, on average, worse for small heads. To ensure that our results were not biased by head size, we calculated individual 'sensitivity maps' ('mne.sensitivity_map' function in MNE-python), which estimate how well each source is sampled by a sensor array (i.e., planar gradiometers). The mean sensitivity in a label was then used as a nuisance variable in our regression analysis.

##### Statistical analyses

Statistical analyses were performed using 'scipy' and 'numpy' Python libraries and STATISTICA-13 software (TIBCO Statistica, v. 13.4.0.14, TIBCO Software Inc, USA). To estimate the group differences in psychometrical variables, head positions, epoch timing, or age, the unpaired t-test or nonparametric tests (Mann Whitney U test, Kruskal-Wallis one-way ANOVA) were applied in case of Gaussian and non-Gaussian distributions, respectively.

To estimate the rank order stability of the spectral slopes across experimental conditions (EO vs. EC), we calculated intraclass correlation coefficients (ICC). We used the ICC version where a fixed set of k 'raters' (two experimental conditions) 'rate' each target (the slope coefficients), while reliability is calculated for the average of k ratings (16).

To estimate group differences in the spectral slope, ANCOVA with factors Group (TD, $ASD_{>85}$, $ASD_{<85}$), Condition (EO, EC), and Age as a covariate was used. Age has been centered (i.e., the group mean has been subtracted from each individual value) before entering it in the analysis (17).

To analyze group differences in the mean periodic alpha and beta power, the power values were log10 transformed to normalize the distributions and ANOVAs with factor Group were used separately for each band and condition.

The relationship between the spectral slope and IQ or age was estimated using Spearman rank order correlations. Non-parametric tests were preferred due to their higher robustness against occasional outliers. To estimate the relationship between the spectral slope and IQ while controlling for nuisance variables (background noise, sensitivity, spectral power), we calculated partial Spearman correlations. Fisher's Z test was used to estimate probability of the difference between Spearman correlation coefficients.

The Benjamini-Hochberg version of False Discovery Rate (FDR) correction for multiple comparisons was applied to the uncorrected p-values of univariate statistical tests applied in the 448 cortical labels.

### SUPPLEMENTARY RESULTS

#### Effect of background noise and motion correction on the spectral slope and high-frequency power

There are two main sources of non-biological (instrumental) noise that may confound spectral power and slope estimation in the high frequency range.

One is the background ('empty room') noise. A high power level of spectrally flat ('white') background noise (Figure 1 in the main manuscript) may result in flattening of the spectral slope measured in the high frequency range. To estimate the background noise, we calculated the mean over all gradiometers power of the 'empty room' signal in the 35-45 Hz range for each subject's dataset.

The other is the noise introduced by the head motion correction procedure. This noise is proportional to the transformation distance (18). To estimate the noise associated with the motion correction procedure, for each subject we calculated the mean transformation distance (displacement of the 'head origin' from the initial position) during the respective condition.

Table S1 shows the correlation of the instrumental noise with the averaged over all cortical labels 35-45 Hz power estimated with sLoreta and the averaged over all cortical labels spectral slope coefficients estimated with the two source localization methods (LCMV beamformer and sLoreta). The correlations were calculated for the pooled sample of participants (N=98) in the EO condition, for which data from all participants were available.

Table S1. Spearman correlations of the 35-45 Hz mean spectral power and slope (Eyes Open condition) with instrumental noise in the pooled group of participants

| | Mean empty room noise | Mean transformation distance |
|---|---|---|
| *Mean 35-45 Hz power* | | |
| *sLoreta* | $R_{(98)}$=0.16, p=0.12 | **$R_{(98)}$=0.23, p=0.02** |
| *Mean slope coefficient* | | |
| *sLoreta* | **$R_{(98)}$=0.33, p=0.0011** | $R_{(98)}$=0.07, p=0.52 |
| *LCMV beamformer* | **$R_{(98)}$=0.24, p=0.016** | $R_{(98)}$=-0.02, p=0.85 |

LCMV, Linearly Constrained Minimum Variance; sLoreta, standardized Low Resolution Brain Electromagnetic Tomography.

As expected, a higher magnitude of the empty room noise correlated with a flatter spectral slope, estimated with either beamformer or sLoreta approaches. The transformation distance, however, did not affect the grand average spectral slope in a systematic way. The correlations of the transformation distance with spectral slopes estimated in the 448 individual labels were also all not significant (only 1 of 448 correlations for the 'sLoreta-estimated' slopes and 5 of 448 correlations for the 'beamformer-estimated' slopes displayed 0.01<p's<0.05, uncorrected for multiple comparisons).

The grand average 35-45 Hz spectral power correlated positively with the mean transformation distance. Although the correlation was moderate ($R_{(98)}$=0.23, p=0.02), inspection of the correlations in the individual labels revealed that they were highest (R~0.5) in the regions most distant from the cranial muscles (caudal midfrontal, paracentral, precentral, postcentral), i.e., in the areas where instrumental noise is the main source of MEG signal contamination.

Considering possible contribution of instrumental noise and myogenic artifacts to the absolute high-frequency spectral power, we suggested that analysis of the group differences in the absolute high-frequency power or 1/f intercept might produce unreliable results. We have therefore chosen not to analyze these parameters.

To ensure that the instrumental noise does not significantly contribute to the observed correlation between the spectral slope and IQ in children with ASD (Figure 7 in the main manuscript), we used the mean empty room noise power in 35-45 Hz range and transformation distance as nuisance variables while calculating partial Spearman correlations between mean spectral slope and IQ in children with ASD. After accounting for these confounding variables, the correlations between the mean spectral slope and IQ remained significant (EO: N=49, $R_{partial}$=-0.44, p=0.002; EC: N=38, $R_{partial}$=-0.38, p=0.02).

**Putative contribution of myogenic artifacts to the 35-45 Hz spectral slope and its group differences**

Muscle activity is the major source of high-frequency signals recorded by surface EEG (19) and is a highly plausible source of MEG artifacts at frequencies >20 Hz (15). Using a semiautomatic procedure for detection of myogenic artifacts (‘annotate_muscle_zscore’ function in MNE-python), we excluded epochs contaminated by *phasic* bursts of myogenic activity from our analyses. Still, *tonic* muscle activity may strongly contaminate the MEG signal recorded by planar gradiometers positioned in the vicinity of the cranial muscles. The power spectrum of the motor units’ activity is broad and is usually concentrated above 50 Hz (20). Therefore, adding this activity to the MEG signal could potentially flatten the slope of the aperiodic component estimated in the 35-45 Hz frequency range.

Beamformers work as spatial filters that reconstruct activity from sources while suppressing interferences from all other sources, including those of muscle artifacts (13, 21). As beamformers reduce the myogenic contribution in the high frequency part of the spectrum, we expected that the spectral slopes estimated with them will be more negative than those estimated using Minimum-Norm-Estimates based methods, such as sLoreta (7). The difference will furthermore be greatest in the regions closest to the sources of myogenic artifacts (i.e., at frontal, temporal, and occipital cortical areas). We also reasoned that if the group differences (TD, $ASD_{>85}$, $ASD_{<85}$) in the mean spectral slope of the aperiodic activity were driven by myogenic artifacts, they would be greater in sLoreta, as compared to beamformer, generated source estimates.

As expected, the local spectral slope coefficients were more negative for the spectra estimated using the LCMV beamformer than for those estimated with sLoreta (Figure S1A,B). Moreover, the differences in the slopes obtained with the two methods were greatest in the frontal, temporal, and occipital regions (Figure S1C).

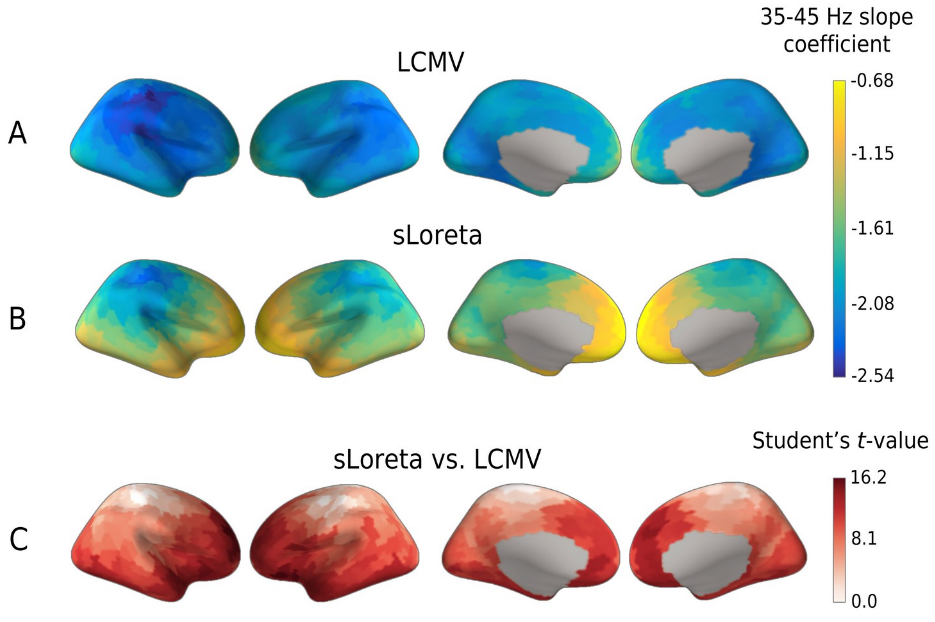


Figure S1. Inflated brain views of the spatial distribution of grand average (N=98) 35-45 Hz spectral slope coefficients of the aperiodic component estimated for the

Eyes Open condition using LCMV beamformer (A) and sLoreta (B). T-values (C) show the difference in the slopes estimated by the two methods (FDR corrected at $p<0.05$). Note that LCMV beamformer yields more negative slopes than sLoreta, but that the difference between the two methods is not significant in the regions most distant from the sources of myogenic artifacts (white spots at tops of brains). LCMV, Linearly Constrained Minimum Variance; sLoreta, standardized Low Resolution Brain Electromagnetic Tomography; FDR, False Discovery Rate.

Next, we analyzed the group differences in the mean (over cortical labels) spectral slopes in the EO condition – for which the MEG data were available in all participants – using 1) beamformer and 2) sLoreta approaches to source estimation. In both cases the ANCOVA with factors Group and Age revealed a significant effect of Group, but the statistical significance and size of this effect were higher in case of the LCMV beamformer ($F_{(2,94)}=8.4$, $p=0.00045$, $\eta^2=0.15$) than sLoreta ($F_{(2,94)}=3.6$, $p=0.032$, $\eta^2=0.07$). The differences in the mean spectral slope between $ASD_{<85}$ and other experimental groups were greater for the LCMV beamformer (Figure S2A) than sLoreta (Figure S2B).

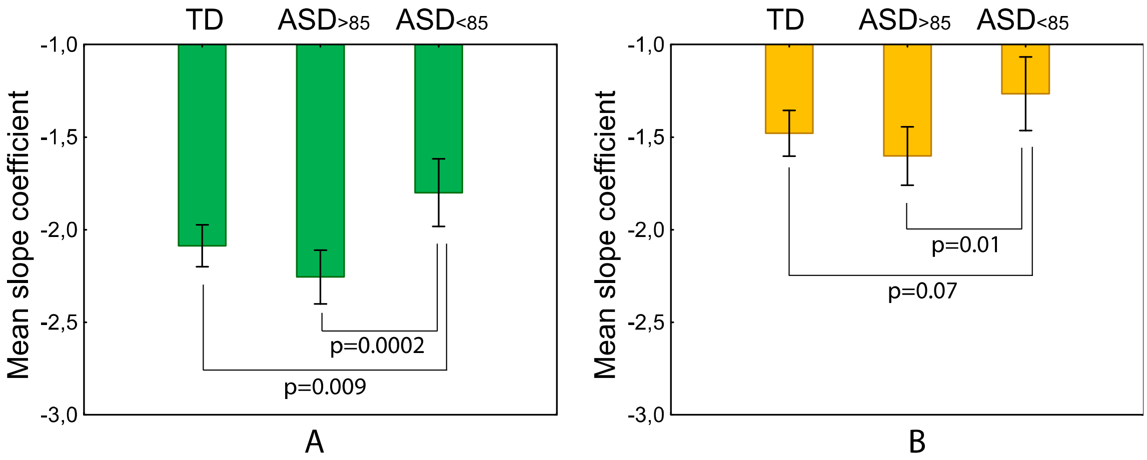


Figure S2. Mean spectral slope calculated in the three groups of participants using LCMV beamformer (A) and sLoreta (B) source estimation methods. Vertical bars denote 0.95 confidence intervals. Note that the group differences are greater in case of the LCMV beamformer, which is a method that is understood to produce source estimates that are less affected by myogenic activity, as compared to sLoreta. The group differences we report in the main text (derived from the LCMV estimates) are therefore unlikely to be explained by a group difference in the amount of myogenic activity. TD, typically developing children; ASD, autism spectrum disorders; $ASD_{>85}$, children with ASD and Mental Processing Index above 85; $ASD_{<85}$, children with ASD and Mental Processing Index below 85; LCMV, Linearly Constrained Minimum Variance; sLoreta, standardized Low Resolution Brain Electromagnetic Tomography.

Inspection of Figure S1 shows that, despite more negative slopes, the distributions of the slope coefficients estimated with LCMV beamformer is similar to that for the sLoreta: the LCMV-based slope coefficients are still less negative at the areas susceptible to myogenic contamination. Therefore, some residual contribution of myogenic artifacts to the high frequency activity, estimated with LCMV beamformers, could not be excluded. Nevertheless, the results presented in Figure 8 of the main manuscript demonstrate that the majority of significant correlations of the local spectral slopes with MPI IQ are found in the regions relatively distant from the cranial muscles. The full list of the cortical labels where the significant correlations survived the FDR correction is presented in Table S2.

In general, this pattern of results suggests against the ‘myogenic nature’ of the observed group differences in the mean spectral slope.

Table S2. The list of cortical labels[a] where the False Discovery Rate corrected partial Spearman correlations between the spectral slope and MPI IQ (controlling for age and sensitivity in the label) were significant in children with autism spectrum disorders

| Label name | Partial Spearman R | P-value |
|---|---|---|
| *Right hemisphere* | | |
| bankssts_2-rh | -0.5 | 0.016 |
| bankssts_3-rh | -0.51 | 0.015 |
| inferiorparietal_1-rh | -0.46 | 0.022 |
| inferiorparietal_3-rh | -0.52 | 0.015 |
| inferiorparietal_4-rh | -0.59 | 0.005 |
| insula_1-rh | -0.37 | 0.05 |
| insula_7-rh | -0.39 | 0.044 |
| lateralorbitofrontal_3-rh | -0.39 | 0.044 |
| middletemporal_4-rh | -0.39 | 0.044 |
| paracentral_4-rh | -0.51 | 0.015 |
| paracentral_5-rh | -0.44 | 0.027 |
| paracentral_6-rh | -0.43 | 0.028 |
| parsopercularis_4-rh | -0.38 | 0.046 |
| parsorbitalis_1-rh | -0.43 | 0.028 |
| parstriangularis_1-rh | -0.42 | 0.029 |
| parstriangularis_3-rh | -0.38 | 0.049 |
| postcentral_3-rh | -0.37 | 0.049 |
| postcentral_4-rh | -0.45 | 0.027 |
| postcentral_7-rh | -0.42 | 0.029 |
| postcentral_8-rh | -0.44 | 0.027 |
| posteriorcingulate_2-rh | -0.39 | 0.045 |
| posteriorcingulate_4-rh | -0.52 | 0.015 |
| precentral_10-rh | -0.43 | 0.028 |
| precentral_11-rh | -0.47 | 0.019 |
| precentral_12-rh | -0.39 | 0.045 |
| precentral_6-rh | -0.4 | 0.041 |
| precentral_7-rh | -0.47 | 0.022 |
| precentral_9-rh | -0.39 | 0.044 |
| precuneus_10-rh | -0.37 | 0.049 |
| rostralanteriorcingulate_2-rh | -0.4 | 0.043 |
| rostralmiddlefrontal_10-rh | -0.45 | 0.026 |
| rostralmiddlefrontal_3-rh | -0.39 | 0.045 |
| rostralmiddlefrontal_5-rh | -0.39 | 0.044 |
| rostralmiddlefrontal_6-rh | -0.39 | 0.044 |
| rostralmiddlefrontal_8-rh | -0.4 | 0.043 |
| superiorfrontal_1-rh | -0.39 | 0.044 |
| superiorfrontal_2-rh | -0.49 | 0.019 |
| superiorfrontal_3-rh | -0.45 | 0.027 |
| superiortemporal_1-rh | -0.38 | 0.049 |
| superiortemporal_2-rh | -0.42 | 0.031 |
| supramarginal_1-rh | -0.45 | 0.026 |
| supramarginal_2-rh | -0.44 | 0.027 |
| supramarginal_3-rh | -0.47 | 0.019 |
| supramarginal_4-rh | -0.44 | 0.027 |
| supramarginal_5-rh | -0.42 | 0.03 |
| supramarginal_6-rh | -0.43 | 0.028 |
| supramarginal_7-rh | -0.44 | 0.027 |
| supramarginal_8-rh | -0.54 | 0.015 |
| supramarginal_9-rh | -0.47 | 0.019 |
| *Left hemisphere* | | |
| cuneus_1-lh | -0.38 | 0.048 |
| cuneus_2-lh | -0.4 | 0.043 |

| | | |
|---|---|---|
| inferiorparietal_2-lh | -0.44 | 0.027 |
| insula_1-lh | -0.42 | 0.029 |
| insula_6-lh | -0.4 | 0.041 |
| lateraloccipital_3-lh | -0.51 | 0.015 |
| parsopercularis_1-lh | -0.38 | 0.046 |
| parstriangularis_2-lh | -0.48 | 0.019 |
| parstriangularis_3-lh | -0.4 | 0.043 |
| pericalcarine_1-lh | -0.4 | 0.041 |
| pericalcarine_2-lh | -0.43 | 0.028 |
| postcentral_11-lh | -0.44 | 0.027 |
| postcentral_12-lh | -0.38 | 0.048 |
| postcentral_13-lh | -0.41 | 0.036 |
| postcentral_14-lh | -0.38 | 0.049 |
| precentral_11-lh | -0.4 | 0.043 |
| precentral_12-lh | -0.38 | 0.049 |
| precentral_8-lh | -0.42 | 0.031 |
| rostralanteriorcingulate_2-lh | -0.44 | 0.028 |
| rostralmiddlefrontal_1-lh | -0.4 | 0.041 |
| rostralmiddlefrontal_10-lh | -0.42 | 0.029 |
| rostralmiddlefrontal_4-lh | -0.48 | 0.019 |
| rostralmiddlefrontal_5-lh | -0.44 | 0.027 |
| rostralmiddlefrontal_6-lh | -0.43 | 0.028 |
| rostralmiddlefrontal_7-lh | -0.43 | 0.028 |
| rostralmiddlefrontal_8-lh | -0.49 | 0.019 |
| rostralmiddlefrontal_9-lh | -0.48 | 0.019 |
| superiorfrontal_2-lh | -0.45 | 0.026 |
| superiorfrontal_3-lh | -0.49 | 0.019 |
| superiorfrontal_4-lh | -0.42 | 0.029 |
| superiorparietal_10-lh | -0.39 | 0.045 |
| superiortemporal_11-lh | -0.38 | 0.049 |
| superiortemporal_2-lh | -0.38 | 0.049 |
| superiortemporal_4-lh | -0.37 | 0.05 |
| supramarginal_1-lh | -0.46 | 0.022 |
| supramarginal_2-lh | -0.53 | 0.015 |
| supramarginal_3-lh | -0.46 | 0.022 |
| transversetemporal_2-lh | -0.43 | 0.028 |

[a] Parcellation into 448 cortical similar-size labels was performed as described by Khan et al. (3).

MPI IQ, Mental Processing Index.

#### Estimation of the spectral slope with FOOOF – 'Fitting Oscillations One Over F'

To estimate the slope of aperiodic component of the spectra in the 2-45 Hz range with FOOOF (14) (hereinafter referred to as the '2-45 Hz spectral slope'), we first filtered the raw signal between 1-47 Hz. The other steps of the source localization and spectral estimation were the same as for the 35-45 Hz spectral slope. For spectrum parameterization in the 2-45 Hz range we used the FOOOF Python package (https://pypi.python.org/pypi/fooof/; v.1.0.0). We extracted aperiodic components from each of the 448 labels' power spectrum with the option 'fixed' (no knee); other parameters were set to default. By multiplying by -1, the scaling exponent provided by FOOOF was converted into the linear slope coefficient on a logarithmic scale. We then calculated the mean (over cortical labels) 2-45 Hz spectral slope in the same way as we did for the slope in the 35-45 Hz range.

Results of ANCOVA with factors Group (TD, $ASD_{>85}$, $ASD_{<85}$), Condition (EO, EC), and Age are shown in Table S3. Unlike the spectral slope directly measured in the 35-45 Hz range, the slope estimated in the broad frequency range (2-45 Hz) using FOOOF did not reveal group differences (Effect of Group: $F_{(2,80)}=0.72$, $p=0.49$, $\eta^2=0.018$; Table S3).

Table S3. Results of GLM analysis for the 2-45 Hz spectral slope estimated with FOOOF: effects of Group (TD, $ASD_{>85}$, $ASD_{<85}$), Condition (Eyes Closed, Eyes Open), and Age

| rmANOVA effect | $F_{(df)}$, p, $\eta^2$ |
|---|---|
| Group | $F_{(2,79)}=0.69$, $p=0.50$, $\eta^2=0.017$ |
| Age | $F_{(1,79)}=29.67$, $p=1e-6$, $\eta^2=0.27$ |
| Condition | $F_{(1,79)}=16.42$, $p=0.0001$, $\eta^2=0.17$ |
| Condition x Age | $F_{(1,79)}=1.51$, $p=0.22$, $\eta^2=0.019$ |
| Condition x Group | $F_{(2,79)}=0.56$, $p=0.57$, $\eta^2=0.014$ |

GLM, General Linear Model; TD, typically developing children; ASD, autism spectrum disorders; $ASD_{>85}$, children with ASD and Mental Processing Index above 85; $ASD_{<85}$, children with ASD and Mental Processing Index below 85; rmANOVA, repeated measures analysis of variance.

Although the difference in results obtained using the two methods of the slope estimation requires further investigation, we believe that, in our study, the spectral slope might not be optimally estimated in the 2-45 Hz range. *Firstly*, a poorly detectable spectral ‘knees’ might present in the data (9, 22) and vary among participants and cortical locations. Since detection of a ‘knee’ in the noisy data and with 1 Hz frequency resolution is problematic, we used ‘no knee’ approach, which could distort estimation of aperiodic activity. *Secondly*, while the FOOOF relies on the assumption that all oscillation peaks are lying within the fitting range (i.e., in our case 2-45 Hz), this might not be the case in all our participants, especially in those with ASD, in whom delta oscillations are increased depending on the severity of their condition (23-25). This might seriously compromise the results, as discussed in a recent paper by Gerster et al (10). When fitting and removing the oscillation peaks seems unfeasible, these authors recommend to estimate the slope of aperiodic component (1/f exponent) at high frequencies (10). *Thirdly,* the source estimation of the low-amplitude high-frequency activity with LCMV beamformers may be imprecise when estimates are based on relatively broadband (2-45 Hz) data. This because beamformer weights that are computed from broadband data are inherently biased towards resolving higher-amplitude low-frequency brain activity (26).
